# Molecular Epidemiology of Trypanosomes in Tsetse Flies in the Livestock-wildlife-Human interface of Eastern Zambia

**DOI:** 10.1101/2024.07.16.24310474

**Authors:** Evans Chikandi, Humphrey Simukoko, Martin Simuunza, Ndonyo Likwa, Lad Slav Moonga, Kalaluka Mbumwae, Mukubesa Andrew, Joseph Ndebe, Queen Suzan Midzi, Wezi Kachinda

## Abstract

**Introduction:** Trypanosomiasis is caused by several species of trypanosomes. The disease is endemic in Eastern Zambia, particularly in the Luangwa River valley. There is a significant threat as many people are at risk of the infection. The disease is classified as a neglected tropical disease. Previously, Trypanosome epidemiological studies in this area focused on using low-sensitive parasitological diagnostic tools to determine the prevalence in animals and some few investigations on trypanosomiasis infection rates in tsetse flies. Livestock production, crop production and jobs in the tourism sector are the primary sources of livelihood in the south Luangwa national park. The South Luangwa National Park settlement town of Mfuwe is an economically and ecologically important area. The presence of trypanosomiasis repetitively threatens tourism and livelihoods, hence the forthcoming need to clarify the trypanosomiasis situation and to advocate the best tsetse and trypanosomiasis control measures in the area. These threats negatively affect the country’s Forex because fewer tourists are visiting.

**Material and methods:** A cross-sectional study that was conducted in Mfuwe of the Eastern Province of Zambia to determine trypanosome infections in tsetse flies. Trapping of the tsetse flies involved the use of epsilon traps and black screen fly rounds in four different vegetation zones, namely open savannah, closed savannah, Montane and riverine. Diagnosis of trypanosome infections in trapped tsetse flies was achieved using a nested PCR which employed the use of two sets of primers targeting the ITS genes.

**Results:** Two hundred twenty-three (223) tsetse flies were captured, and only 213 were processed for laboratory analysis. The highest catches from the areas selected were (153) of Tsetse flies in montane, followed by areas near the open forest (lagoons), (48) whereas the lowest catches was recorded in the riverine areas with 22 catches. The species found in these areas were *Glossina morsitans morsitans* and *Glossina palpalis,* but *Glossina brevipalpis* were not found,. The overall prevalence using a nested PCR of the identified trypanosome infection rates in tsetse flies was 18.3% (p < 0.001). The difference in prevalence between sampling areas were significant. Statistically significant differences (p < 0.001) were observed when the prevalence of trypanosome infections was compared by season. The Trypanosoma species found were *T.congolense, T.vivax, T.brucei* and *T.theileri*.

**Conclusions:** The results of this study showed that location has a significant contribution to the trypanosome infection rates in the tsetse vector at the Wildlife-Livestock-human interface of the Luangwa valley. The results are essential for designing community-wide tsetse and trypanosomiasis control interventions and planning sustainable regimes for mitigating the burden of trypanosomiasis.

## Introduction/Background

Trypanosomiasis is caused by several species of trypanosomes including Trypanosoma *congolense, T.vivax, T.godfrey, T.Simiae* and T.brucei (Laohasinnarong *et al.,* 2015). The main tsetse fly species present at the wildlife-Livestock-human interface of the Luangwa River valley of eastern Zambia are *Glossina pallidipes austen* and *Glossina morsitans morsitans* which are responsible for the trypanosomiasis transmission dynamics. Trypanosomiasis is considered as an neglected tropical disease associated with an agriculture loss of up to US$4.5 billion per year, with as many as three million cattle dying each year worldwide (Sutherland *et al.,* 2015). Trypanosomiasis is endemic in Eastern Zambia, particularly in the Luangwa River valley where there is a large tsetse fly population due to abundance of wildlife in game parks and domestic animals in the surrounding settlement areas. There is a significant threat as many people and Livestock are at risk of the infection. About 30%-50% of AAT infections in eastern Zambia are caused by *T. congolense* (Simukoko *et al*., 2007; Laohasinnarong *et al.,* 2015). Tsetse flies’ survival is highly dependent on the presence of suitable habitat and hosts. Habitat fragmentation affects tsetse fly distribution since tsetse flies depend on vegetation for resting and reproduction (Mweempwa *et al.,* 2015). The vegetation in Eastern Zambia has changed over the years mainly due to human interference and this has affected the tsetse distribution patterns and therefore the trypanosomiasis epidemiological situation in the area has also been affected (Laohasinnarong *et al.,* 2015).

In the Luangwa Valley of Eastern Zambia, the main tsetse fly hosts are the wild animals and opportunistically Livestock and humans. Livestock and humans live along the edges of the South Luangwa National Park but Livestock and humans also constantly infiltrate the national park. Livestock infiltrates the fringes of the national park mainly in search of grazing pastures, while humans infiltrates the national park for activities mainly related to tourism, hunting, and firewood fetching and fishing. The penetration of Livestock and humans on the fringe and deep in the national park puts both humans and animals at risk of Trypanosoma infections. The constant movements over the years of Livestock, humans, wildlife and vehicles to and from the national park affects the tsetse flies in the sylvatic area, which cross into the domestic areas and vice-versa, thereby affecting the trypanosomiasis epidemiological dynamics (Laohasinnarong *et al*., 2015).

The South Luangwa National Park is made up of a natural semi-arid ecosystem providing a home for a variety of animal species and vegetation. Livestock, crop production and jobs in the tourism sector are the primary sources of livelihood. The presence of trypanosomiasis constantly threatens tourism and livelihoods, hence the imminent need to suggest the best tsetse and trypanosomiasis control measures in the area. Previously, molecular diagnostic tools had not been used to investigate tsetse fly infection rates and to elucidate the trypanosomiasis epidemiological setup on the wildlife-Livestock-human interface of the Luangwa River valley of Eastern Zambia. The study used a molecular diagnostic tool (nested PCR) to investigate trypanosomiasis infection rates in tsetse flies.

These step-up areas have different influences on the abundance of tsetse fly. Further, its proximity to the protected areas, the study had high interaction of wild, domestic animals, and humans which increases circulation of trypanosomes through bites from tsetse flies, making trypanosomiasis endemic in the area. Trypanosomiasis is of public health importance in humans and is also the significant constraint for Livestock productivity. The progression of trypanosomiasis in animals varies depending on the factors associated with the host and the parasite.

Diagnosis relies on laboratory techniques that confirm the presence in the blood of trypanosomes or the presence of anti-trypanosomal antibodies and also molecular techniques. Molecular techniques such as polymerase chain reaction (PCR) have significantly improved the sensitivity and accuracy of trypanosome diagnosis compared to the traditional parasitological methods (Mulenga *et al*., 2021). Molecular tests differentiate between trypanosome species and subspecies using specific primers (Mulenga *et al*., 2021). Molecular identification of trypanosomes by the Polymerase Chain Reaction (PCR) in tsetse flies is specific and robust in showing no reaction with non-target trypanosome DNA or a massive excess of host DNA. Thus, PCR remains the right diagnostic tool for the accurately identifying of trypanosome species and subspecies.

## Material and methods

The research took place in Mfuwe, which is situated in the Eastern Province of Zambia. This region falls between latitudes 10° and 15° S 0.478’ and longitude 30° and 33°47.30’ east. The Eastern Province shares its borders with Malawi to the east and Mozambique to the south, and it accounts for approximately 9% of Zambia’s total territory, covering an area of 69,000 km2. The province comprises various districts, including Chipata, Chama, Lundazi, Chadiza, Mambwe, Nyimba, Katete, Petauke, Sinda, and Luangwa, with this particular study being conducted in Mfuwe, which is located in the Mambwe district. The area experiences three distinct seasons: the warm, wet season (from November to April), the cool, dry season (from May to August), and the hot, dry season (from September to October). The average annual rainfall is about 1000 mm, with most of the rains occurring between December and March (Simukoko *et al.,* 2007). The region experiences an average temperature ranging between 18 °C and 30 °C. The South Luangwa National Park is a natural semi-arid ecosystem that provides a suitable habitat for various animal species, vegetation, and favorable temperature variations and rainfall, which facilitate the survival of vectors and parasitic organisms.

To conduct this research, a cross-sectional study design was employed. Two sampling periods were carried out during the start of the wet and the beginning of the dry season in each habitat type. This allowed for a comparison of the prevalence of trypanosome infections in tsetse flies over the sampling intervals. The vegetation in Mfuwe consists of four broad types, including plateau miombo woodland, valley miombo woodland, scrub miombo woodland, and scrub and woodland mopane. The study area was chosen due to its proximity to areas where game animals are located and go for drinking water.

South Luangwa National Park has the highest diversity of tsetse fly species in Zambia (Signaboubo. *et al*., 2019). The research conducted entomological surveys in Mfuwe, where tsetse flies were trapped from game areas for five consecutive days using odour-baited traps and black screens fly rounds placed in suitable tsetse fly habitats. Nine traps were deployed, with three placed in each habitat, and the geographical coordinates of each trap were recorded using GPS. Tsetse flies were collected twice a day at 11:00 hours and 16:00 hours.

Tsetse population monitoring was carried out by black screen fly rounds along two sub-transects at each site, which meandered through various ecotones and traps. Each sub-transect was divided into sectors of 250 m in length and traversed at an average of two times per day during the five days of sampling.

The species and sex of the trapped flies were identified using morphological criteria, and young flies that had never taken a blood meal were excluded from the sample. Non-teneral flies were dissected, and the body parts were separately transferred into Eppendorf tubes and preserved at - 20°C until further analysis. DNA was extracted from the tsetse flies using the Quick-gDNA blood mini prep kit and stored at −20 °C until further analysis.

A nested PCR was employed using two sets of primers comprising outer forward (5′-GAT TAC GTC CCT GCC ATT TG-3′) and reverse (5′-TTG TTC GCT ATC GGT CTT CC-3′) primers, and inner forward (5′-GGA AGC AAA AGT CGT AAC AAG G-3′) and reverse (5′-TGT TTT CTT TTC CTC CGC TG-3′) primers targeted the Internal Transcribed Spacers (ITS). The primary PCR (nPCR) reaction was conducted in a 12.5 μl reaction volume comprising 10 μl master mix (Quick-Load Taq 2× Master Mix, New England BioLabs Inc., Ipswich, MA, USA), 1 μl DNA template, 6.30 μl nuclease-free water and 0.8 μM of each first-round primer.

The secondary PCR amplification was performed using primers at the same concentration as the first round, 1 μl of the PCR products from the first-round was used as a template and 4.75 μl of n nuclease-free water. Amplification conditions involved an initial denaturation step at 98 °C for 30 sec followed by 45 cycles of denaturation at 98 °C for one min, an annealing step at 59 °C for one min, then an extension step of 72 °C for two min and a final extension at 72 °C for 2 min. PCR products were loaded on 1% agarose gel stained with Ethidium bromide and visualized on a Gel DocTM (Bio-Rad, Hercules, CA, USA). Additionally, a single-step PCR to amplify the ITS 1 gene to determine and measure the trypanosome infection rates in tsetse flies. Briefly, primers with the following sequences were used: F: 5′-CCG GAA GTT CAC CGA TAT TG-3′ and R: 5′-TTG CTG CGT TCT TCA ACG AA-3′ The reaction was performed in a 25 μl reaction volume containing 12.5 μl of master mix (Quick-Load Taq 2× Master Mix, New England BioLabs Inc., Ipswich, MA, USA) containing DreamTaq DNA polymerase supplied in 2×DreamTaq buffer, 0.4 mM of each of the dATP, dCTP, dGTP and dTTP, and 4 mM MgCl2, 0.2 μM of each of the forward and reverse primers, 6.3 μl nuclease-free water and 5 μl DNA template. The reaction was conducted in a thermocycler (ProFlex PCR system, Applied Biosystems, Foster City, CA, USA) with an initial denaturation step of 98 °C for 3 min, followed by 30 cycles of 98 °C for 30 s, 60 °C for 30 s, 72 °C for 30 s and a final extension step at 72 °C for 2 min finally, I ran an Agar gel electrophoresis to see the bands.

## Data analysis

To estimate the prevalence of infections, frequency and contingency tables in Excel and statistical software were utilized. Contingency tables and logistic regression were used to estimate relative location and seasonal prevalence, and p-values were obtained to assess statistical significance. Pearson’s uncorrected test was employed to test for independence and homogeneity of proportions between the dependent (outcome) and independent (exposure) variables. All p-values for tables with zero values were obtained with the assumption that 0.5 added. The p-values of the results were analyzed (using p < 0.05 as a cut-off value) at 95% confidence intervals to indicate the level of uncertainty around the obtained values.

## RESULTS

The initially planned sample size was 384, but only 223 tsetse flies were captured during the survey. Some of these flies were excluded from analysis due to being teneral, dried up, or missing body parts, likely due to ants attacking them. After selection, only 213 fresh flies remained for laboratory analysis as some were lost during crushing. A number of factors contributed to the failure to reach the planned sample size, including wild animal interference with the traps, human interference, and heavy rains.

Table 1 provided information on measurements and scales for each data variable, while Tables 2 presented the findings on the number of flies captured using traps in nine different locations. The prevalence of trypanosome infections varied significantly between the sampling areas, as shown in Table 3, and was higher in G.M.M than G.P Tables 4. The prevalence of trypanosome infections also varied significantly between sampling seasons, as demonstrated in Tables 2.

**Table 1.** A table showing Variables, indicators of measurements, and scale of measurements for each variable.

| Variable Type | Indicators of Measurements | Scale of Measurements |
| --- | --- | --- |
| The dependent Variable:<br>Prevalence of Trypanosome infection in Tsetse flies | 1. Proportion of Trypanosome infection in Tsetse flies. | Binary |
| Independent Variables:<br>1.The population Size | Number of Tsetse flies in all areas. | Count |
| 2.Population Density | Number of Tsetse flies in a location per square kilometre. | Ordinal |
| 3.Demographic Characteristics of Tsetse Flies:<br>Age<br>Sex | Life span survival in months or years<br>1.Female<br>2, Male | Nominal<br>Binary |
| 4.Seasonal variation<br>Interval<br>Frequency | Weekly<br>Monthly<br>Quarterly<br>Annually | Ordinal |

**Figure 1.**
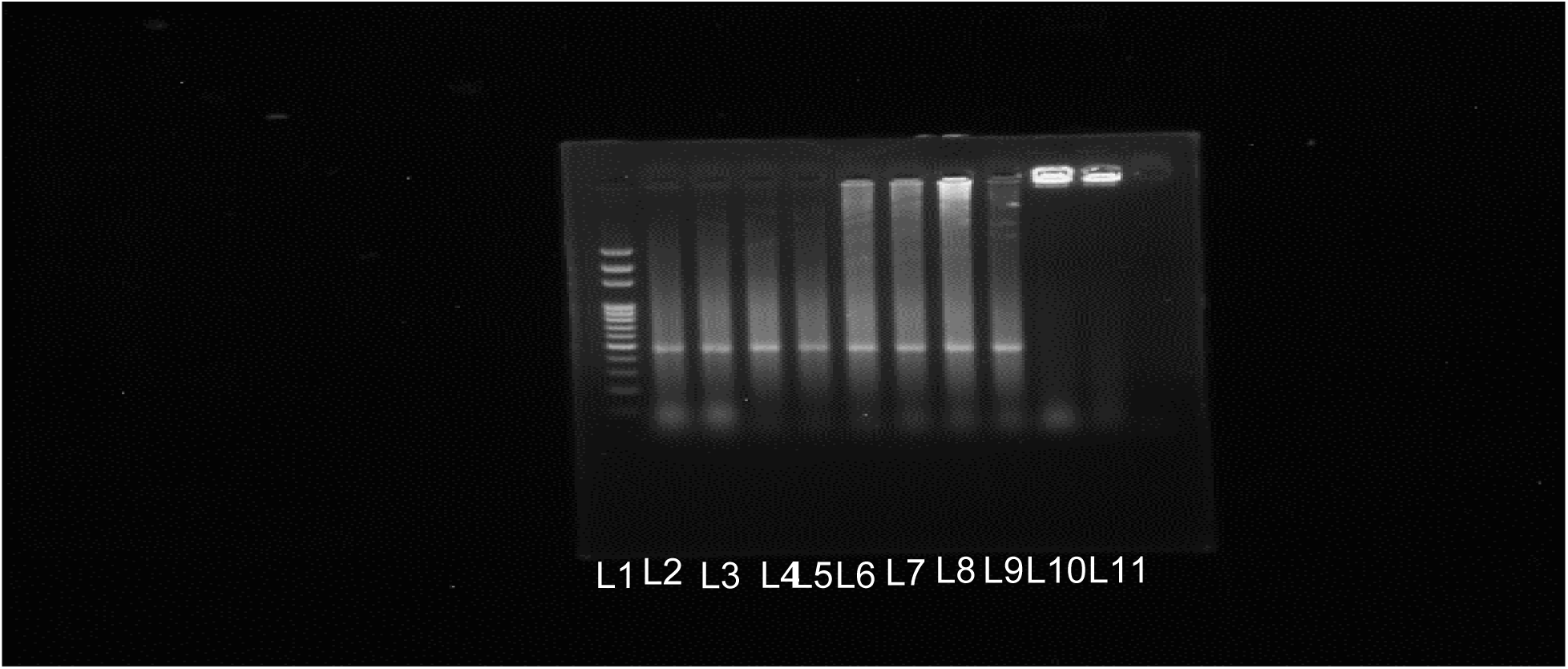
Gel electrophoresis picture of trypanosomes amplified using nPCR. Lane L: 1 100-base pair (bp) DNA ladder; Lanes 11 Negative control and Lanes 2 through to lane 9 are positive samples for T. congolense separated from the pooled samples, and individually run and lane 10 was doubtful. This was just an example of one run for *T. congolense* and other results for *T. vivax* were not shown.

**Table 2.** Prevalence of trypanosome infections in the study population of tsetse by different ecotone by traps deployed and black screen.

| Category | No. of tsetse screened | Prevalence of trypanosomes (%) | P-value |
| --- | --- | --- | --- |
| Open savanna | 99 | 99/213 (21.42) | p <0.001 |
| Montane | 48 | 48/213 (35.40) |  |
| Riverine | 22 | 22/213 (22.7) |  |
| Black screens round | 54 | 54/213 (14.8) |  |
| **TOTAL** | **213** | **18.3** |  |

**Table 3.** Prevalence of trypanosome infections according to different species of Flies in different locations.

| Species | Total No. of | No. of | p-value |
| --- | --- | --- | --- |
|  | Infections | Infection (%) |  |
| T. vivax | 17 | 43.5 | p < 0.001 |
| T. Congolense | 11 | 28.2 |  |
| T. brucei | 08 | 20.5 |  |
| T. Simiae | 00 | 00.0 |  |
| T. theileri | 4 | 10.1 |  |

**Table 4.** Prevalence of Trypanosomes Distribution.

| Species of Tsetse | N | Species of Trypanosomes | Prevalence % | p-value |
| --- | --- | --- | --- | --- |
| G.M.M | 103 | T. vivax 38 | 38/103*100 37% | p < 0.001 |
|  |  | T. congolense 12 | 12/103*100 12% |  |
|  |  | T. brucei 53 | 53/103*100 51% |  |
| G. P | 90 | T. vivax 23 | 23/90*100 26% |  |
|  |  | T. congolense 16 | 16/90*100 18% |  |
|  |  | T. brucei 51 | 51/90*100 57% |  |

Tsetse flies were tested for trypanosome infections using the ITS PCR to determine spatial and temporal variation in prevalence across different tsetse fly species. The overall prevalence of trypanosomes in tsetse flies was 18.3%, but different trends were observed in different areas. Higher levels of destruction were associated with lower prevalence and infection rates of the parasites, as demonstrated by PCR band results at 500Bp. The highest number of captures were observed in Montane (closed forest), followed by areas near lagoons (Open Savannah), while the riverine areas recorded the lowest captures, and the Black screen fly rounds recorded 54 captures, which was part of the montane.

## DISCUSSION

The 18.3% prevalence of trypanosome infections in tsetse flies in the different sampling study locations was established. The study was performed to provide solutions that would have implications on the control of trypanosomiasis in the Livestock-wildlife-human interface of Eastern Zambia. The first objective of this study was to determine the prevalence of trypanosome infection rates in tsetse flies. Also, the variation in the population size and density of tsetse in various vegetation types using molecular techniques. Our study was positioned on the fringes of the South Luangwa National Park located within the Luangwa valley. The wildlife species in this area mainly comprised buffalo (10,061), Impala (7,531), puku (3,626), hippo (2,853), and elephant (2,183) among others (Aerial survey report: Luangwa valley 2012). Buffalos and hippos were found congregating near lagoons in search of water within the Game Management Areas (GMAs). This influenced the prevalence of some species of tsetse flies, as evidenced by their feeding habits. According to the results of the study, the highest tsetse abundance was recorded in montane followed by Open Forest and most minor abundance were recorded in the riverine (Table 3).

Thus, the differences in the tsetse flies trapped may have been due to tsetse feeding preferences and the type of traps used in this study. The main trap used in our study was Epsilon. Biconical traps are more efficient in trapping *G. morsitans* while F3, epsilon and NGU traps are more efficient in trapping *G. brevipalpis* (Kweka *et al.,* 2017). Therefore, the prevalence (18.3%) reflected trypanosome infections in tsetse regardless of the season at the time of sampling. The prevalence level further indicated the potential risk for tourists and pastoral cattle to suffer from trypanosomiasis and spread the infections to other animals through tsetse fly bites. There were a lot of differences in the number of tsetse flies trapped in different study locations where traps were deployed. The reason for variation was the proximity to areas where the wildlife go most for seeking good grazing and drinking areas which favored much the attraction of the flies. Furthermore, the difference in the prevalence was the sensitivity of PCR compared to microscopy in detecting trypanosomiasis. PCR detected both clinical and subclinical infections.

The planned minimum sample size calculated was 384. However, the actual total numbers of tsetse flies trapped in the survey was 223 which were lesser. Moreover, the number of tsetse flies that were analyzed to determine trypanosome prevalence was 213. Likely this could have affected the estimate’s precision and also might have reduced the power of the study. However, this did not affect our results since the calculated p < 0.001 indicated strong data reliability. The low sample size was because some flies were removed because they were teneral flies. Others dried up, while the remaining ones only had heads. The probable reasons for this scenario were mainly due to ants eating up the tsetse flies. Few flies were lost during crushing. Other encounters for failure to get the planned minimum sample size included wild animals disturbing the traps, human primates tempering and bringing down the traps and heavy rains. However, the obtained prevalence was not significantly different to that obtained by other studies (Signaboubo *et al.,* 2021; Weber *et al*., 2019), which might be an indication that the low sample size in our study might not have had a significant effect on the precision of the estimate. (Ngonyoka, *et al*., 2017) in Tanzania reported higher tsetse fly abundance in Acacia-Swampy ecotone and riverine habitats. These results differed from our study results, which showed higher abundance in the montane habitat with lower abundance in the riverine habitat. The discrepancy could be due to the climatic condition in Tanzania which has two rainy seasons. This favored the fly densities. In our study the most trapped tsetse fly species were *G. pallidipes* followed by *G. morsitans morsitans* while no *G. brevipalpis* were trapped. As observed, the variation in the abundance of tsetse flies in the three habitats is influenced by the quality of the vegetation and the abundance of hosts.

In our study, trypanosome infection rates were highest in the montane habitat, while the lowest was recorded in the open forest (habitat). The characteristics of tsetse fly habitats, host-vector interactions and consequently, tsetse infection rates were affected by transformations in land cover (Malele *et al*., 2016). Indeed, the studies demonstrated variations of tsetse fly species abundance and infection rates among different habitats.

It has been observed that the difference in trap efficiency is related to the behavioural differences among the tsetse fly species and varies between different populations of the same species (Malele *et al*., 2015). The closeness to the lagoons and good vegetation had also influenced trap efficiency. Results obtained reinforced by previous studies that the Livestock-human-wildlife interface of Eastern Zambia still has trypanosome infested tsetse flies. Therefore, the study’ results further showed that the tsetse flies circulating in the area and infected with trypanosomes could still pose a high risk of Livestock and human trypanosomiasis. Indeed, according to the results of the study, the highest infection rates were recorded at the end of the wet season in April. This was due to the favorable conditions during the wet season with the highest fly burden.

The study was very significant because it reflected the situation of infection rates in Mfuwe compared to the previously reported research on the prevalence of trypanosomes in tsetse flies. This prevalence was higher than the 5% reported by scholars who used parasitological (microscopy) methods to detect active disease in the same area using blood samples (Simwango, *et al*., 2017).

The prevalence of trypanosome infections was found to be higher during the dry season compared to the wet season. Although the difference between the two sampling seasons was not statistically significant, this outcome was expected because the game tended to graze in tsetse fly-infested areas in the dry season, such as artificial lagoons, while in the wet season, they grazed everywhere due to the availability of grass, resulting in less exposure to tsetse fly bites. Our study revealed a significant variation in the prevalence of trypanosome infections across different study locations. The variation could be attributed to differences in community practices for vector control and livestock movement towards protected areas. In Mfuwe, seasonal livestock movement from one location to another in search of pasture and water is a common occurrence, and these movements were expected to influence the prevalence of trypanosome infection rates.

Furthermore, the movements of settlers with their herds of cattle increases their interaction with wildlife through shared grazing fields, hence the potential for disease transmission between wild and domestic hosts (Simwango *et al*., 2017). Vector control is crucial in areas where cattle are present, particularly in mountainous regions and lodges where the incidence of trypanosomes is high, as well as in open savannas and riverine areas that are located closest to the National Park. Although appropriate habitats for tsetse flies can still be found in all levels of fragmentation, it is anticipated that the degree of fragmentation affects the movements of these insects as they search for food. As many of the wild hosts have disappeared from the study area, tsetse flies have become heavily reliant on domesticated animals for their survival. While domesticated animals are most readily available in highly fragmented landscapes, their distribution varies according to the season. Therefore, tsetse is highly dependent on seasonal changes in their distribution to find their host (Van den Bossche and De Deken, 2006). Further investigation is needed to understand how changes in apparent fly density affect disease transmission. Another abundant wildlife species in Mfuwe is non-human primates, particularly baboons, but their role in trypanosomiasis transmission is poorly understood, so additional research is needed. Non-human primates may also contribute to the transmission of sylvatic to domestic trypanosomes as they move in and out of the game park and the GMA, potentially increasing the distribution of tsetse flies in the area. Vehicular traffic dynamics can also impact the prevalence of tsetse flies in GMAs and settlement areas, as tsetse flies tend to follow moving vehicles.

The prevalence of trypanosome infection rates in tsetse flies is significantly influenced by seasonal and spatial factors compared to other studies. These findings are important for designing community-wide vector and disease control interventions and planning sustainable regimes to reduce the burden of trypanosomiasis. They can also help strengthen trypanosomiasis control measures at the local, regional, and national levels. Although the sample size was lower than calculated, this study established the seasonal and spatial prevalence of trypanosome infections in tsetse flies across various locations in Mfuwe, eastern Zambia. The study found that the prevalence of trypanosome infections was positively influenced by the proximity of the sampling site to wildlife, with a high number of tsetse flies carrying trypanosome infections.

This study demonstrated the circulation of different trypanosome infection rates and factors influencing their occurrence. The geographical distribution of trypanosome species and tsetse flies can be used to guide improved control measures and strengthen trypanosomiasis control at the local, regional, and national levels. The approach presented in this paper may also serve as a basis for rationalizing area-wide tsetse fly control activities, such as identifying areas where tsetse flies are likely to disappear without control measures and areas where control is necessary.

In conclusion, further investigation is required to understand the impact of changes in apparent fly density on disease transmission. The findings from this study are significant for designing effective community-wide vector and disease control interventions and planning sustainable regimes to reduce the burden of trypanosomiasis. They can also help strengthen trypanosomiasis control measures at the local, regional, and national levels, while identifying areas where tsetse fly control is necessary and where it is not, helping to reduce negative impacts on the hospitality business and foreign exchange in Mfuwe and Zambia.

## Data Availability

N/A

## Acknowledgements

The authors would like to acknowledge Zambia wildlife authority in Chilanga and Mfuwe, as well as the following individuals from Chipata Veterinary Office for their technical support: Anthony Chupa, Sikazindu, Kalaluka Mbumwae, Kaluba Andrew Chibango and Michael Mhango.

More acknowledgements go to Africa Centre of Excellence for Infectious Diseases of Humans and Animals (ACEIDHA) for funding my education and my supervisors.

## Institutional Ethical Approval

Institutional ethical approval was acquired from the University of Zambia Biomedical Research Committee (UNZABREC).

## Informed Consent Statement

Prior to the investigations, written informed consent was obtained from all the participants. Conflicts of interest: None declared by the authors.

## Notes

### Competing Interest Statement

The authors have declared no competing interest.

### Funding Statement

The author(s) received no specific funding for this work.

